# A Network Target for Memory Dysfunction Derived from Brain Lesions and Stimulations

**DOI:** 10.64898/2026.03.10.26348082

**Authors:** Calvin W. Howard, Savir Madan, Arun Garimella, Frederic Schaper, Isaiah Kletenik, Marcus C. Ng, Philip Mosley, Jordan Grafman, Rohit Bakshi, Bonnie Glanz, Lisa Fosdick, Amy Johnson, Ryan Colyer, Constantine G. Lyketsos, Mae Morton-Dutton, John Giftakis, Yasin Temel, Rob P.W. Rouhl, Ji H. Ko, Rabea Schmahl, Juan C. Baldermann, Özgür Onur, Pablo Andrade- Montemayor, Veerle Visser-Vandewalle, Jens Kuhn, Maurizio Corbetta, Robert S. Fisher, Thomas Picht, Katharina Faust, Molly Hermiller, Joel Voss, Tanuja Chitnis, Michael K. Kahana, Gwenn S. Smith, Andres Lozano, Shan H. Siddiqi, Andreas Horn, Michael D. Fox

## Abstract

Therapeutic brain stimulation holds promise in treating memory dysfunction, but recent clinical trials have produced heterogenous results. Uncertainty in the neuroanatomical target may contribute to this heterogeneity, with over 13 different brain regions targeted to date. To derive a neuroanatomical target, we studied verbal episodic memory changes across 1247 patients in 12 independent datasets, including patients with focal lesions (n = 985), DBS sites (n = 207), and TMS sites (n = 72). We found lesion and stimulation sites causing verbal memory changes converged on a common brain network. In 3 held-out datasets, connectivity to this convergent memory network explained more variance than connectivity to modality-specific maps or other neuroanatomy previously implicated in memory. In a meta-analysis of 21 prior brain stimulation trials for Alzheimer Disease, overlap between stimulation site and our convergent memory network correlated with trial effect size. In conclusion, we find Lesions, DBS, and TMS sites influencing verbal memory converge upon a single memory network, and this network may inform targeting in memory neuromodulation trials.

## Introduction

Episodic memory dysfunction arising from brain diseases such as Alzheimer’s is a prevalent problem,^1^ and new treatments are needed.^2,3^ Therapeutic brain stimulation can improve other treatment-refractory neurologic and psychiatric symptoms,^4–6^ and has shown some promise in treating episodic memory dysfunction.^7–9^

However, the efficacy of brain stimulation depends on location of stimulation, and the optimal location for modulating episodic memory remains unclear. Trials of transcranial magnetic stimulation (TMS) have targeted at least 10 different brain regions^8–19^ while trials of deep brain stimulation (DBS) have targeted 3 different brain regions.^7,20,21^ Even within a trial, there is often heterogeneity in the location of stimulation across individuals, which has been linked to heterogeneity in clinical response.^20,22,23^ While there are likely many factors that contribute to heterogeneity in brain stimulation outcomes,^24,25^ refining the neuroanatomical target is a modifiable factor that might be used to improve outcomes.

One approach that has proven successful in refining brain stimulation targets is to analyze data sources that allow for causal links between symptoms and human neuroanatomy.^26^ Prior studies have sought to improve our localization of memory based on brain lesions,^27–31^ DBS,^32–35^ or TMS^15,16,36^ datasets, but have yet to combine these modalities. Combining multiple causal sources of information can mitigate confounds inherent to one modality alone and further refine therapeutic targets, a technique termed convergent causal mapping.^26^ This approach was recently used to refine brain stimulation targets for depression and tremor,^37,38^ but to our knowledge this approach has yet to be applied to episodic memory dysfunction. Further, it remains unclear how best to combine different sources of causal information into a single neuroanatomical target.

Here we tested if the effects of lesions, DBS, and TMS upon verbal memory, the primary proxy of episodic memory,^39,40^ converged on a common neuroanatomical network. We developed methods to define the topography of this network and validated our results in held-out lesion, DBS, and TMS datasets. Finally, we explored the potential of this network as a therapeutic stimulation target for Alzheimer Disease by investigating if the effect size of prior stimulation studies for Alzheimer Disease correlated with their target’s overlap with the convergent memory network.

## Results

### Cohort Characteristics

For our discovery dataset, we analyzed data from 1247 patients spanning 12 independent cohorts, selected because they included objective assessments of verbal episodic memory. These 12 cohorts included patients with brain lesions (total n = 985), DBS (total n = 180), and TMS (total n = 82) (**Table 1**). We considered single-subject locations of Alzheimer-related atrophy a type of “lesion” for the present analyses, using previously defined methods.^41,42^ The lesion, DBS, or TMS site for each patient was localized and warped to a common brain space (**Figure 1**).

**Table 1.**
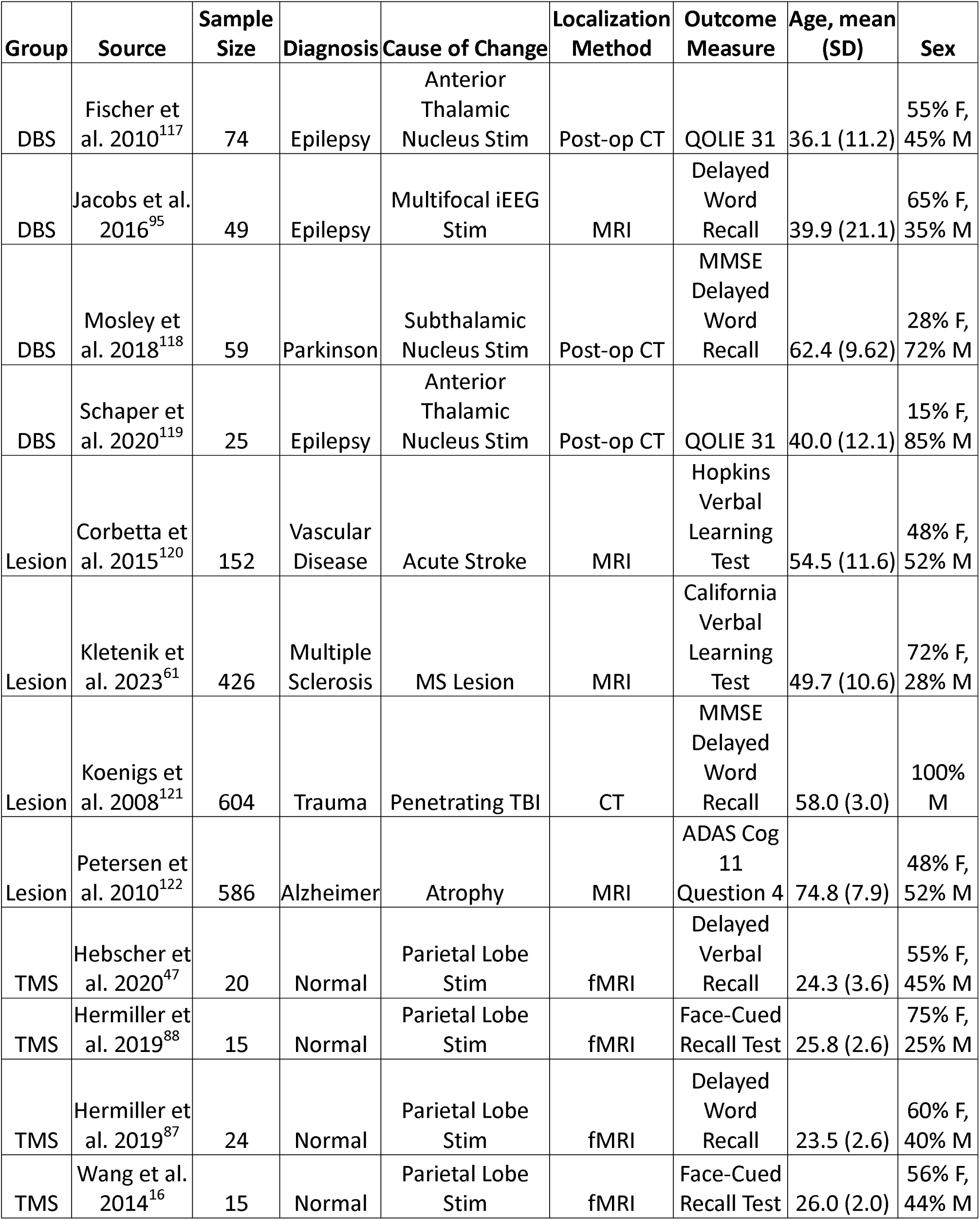
Cohort characteristics.

**Figure 1.**
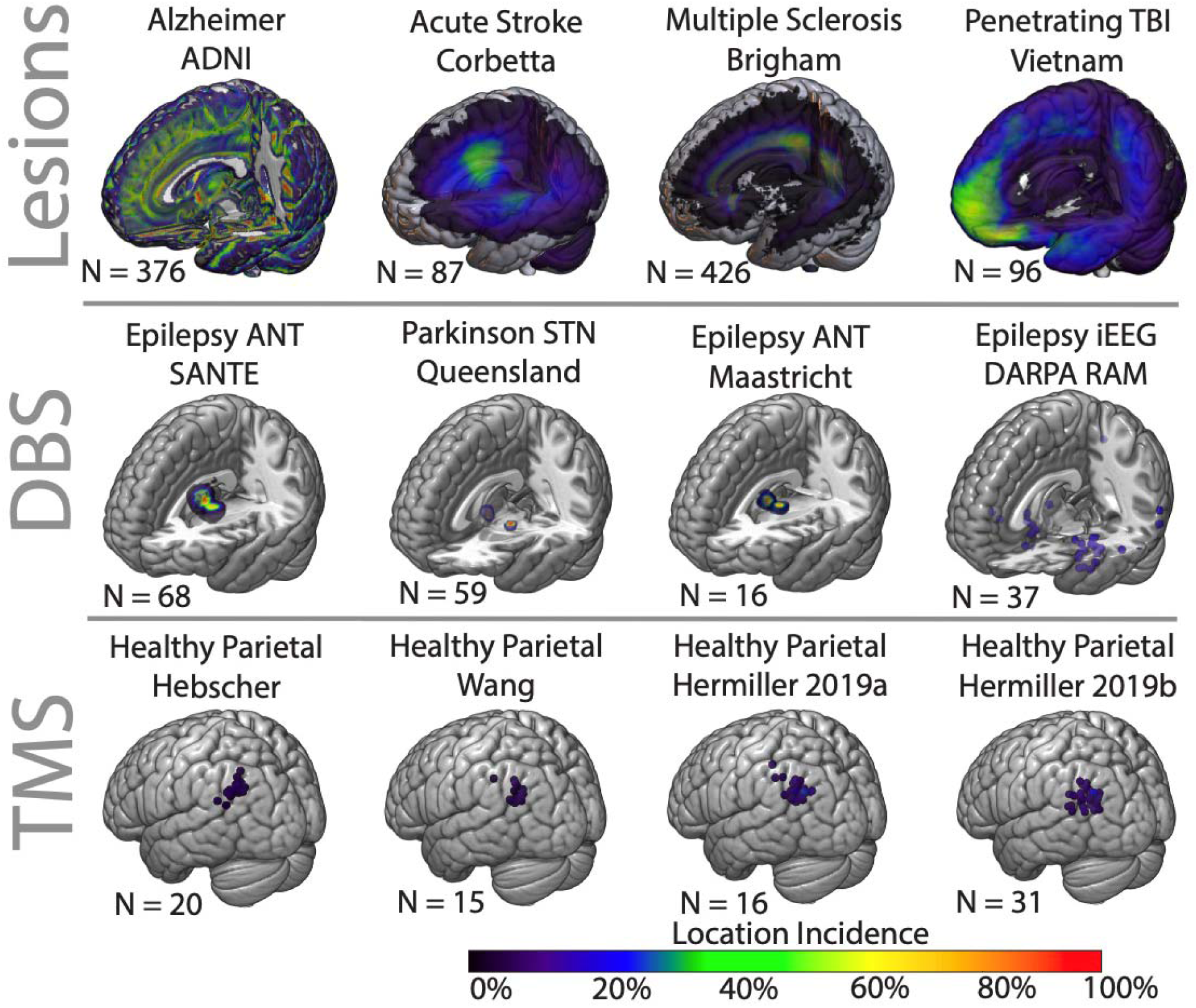
Brain lesions and stimulation sites influencing memory in 1247 patients. Top) Lesion sites influencing memory across 4 datasets, including atrophy In Alzheimer Disease (n = 376), acute strokes (n = 87), multiple sclerosis (n = 426), and penetrating traumatic brain injury (n = 96). Middle) DBS sites influencing memory across 4 datasets, including the anterior nucleus of the thalamus in epilepsy (n = 68, n = 16), the subthalamic nucleus in Parkinson Disease (n = 59), and multifocal iEEG in epilepsy (n = 37). Bottom) TMS sites influencing memory across 4 datasets, including parietal stimulation in healthy controls (n = 20, n = 15, n = 16, n = 31).

### Networks Associated with Memory Changes are Similar Across Cohorts

We calculated the whole-brain connectivity of each patient’s lesion, DBS, or TMS site using established methods and a normative functional connectome derived from 1000 healthy subjects.^43,44^ We identified connections that correlated with verbal episodic memory outcomes in each cohort, resulting in 12 cohort-level memory network maps (**Figure 2**). Within a modality, these networks were more similar than expected by chance (r = 0.53, p = 0.0081) including when assessed separately for lesion cohorts (r = 0.45, p = 0.047), DBS cohorts (r = 0.48, p = 0.026), and TMS cohorts (r = 0.68, p = 0.006). Comparing across modalities, the DBS and TMS cohorts shared similar network topographies and signs (r = 0.43, p = 0.002). However, the lesion cohorts had similar topography but the opposite sign compared to both the DBS cohorts (r = -0.35, p = 0.045) and TMS cohorts (r = -0.38, p = 0.001). These findings were robust to methodological variation and similar using Pearson correlation, Spearman correlation, network cosine similarity, the absolute value of networks, or the squared value of the networks (Supplementary Table 1). These results also depended on network connectivity, as similar convergence across cohorts was not seen with traditional analyses focused on the lesion or stimulation site alone (Rho = 0.04, p = 0.94, Supplementary Figure 1).

**Figure 2.**
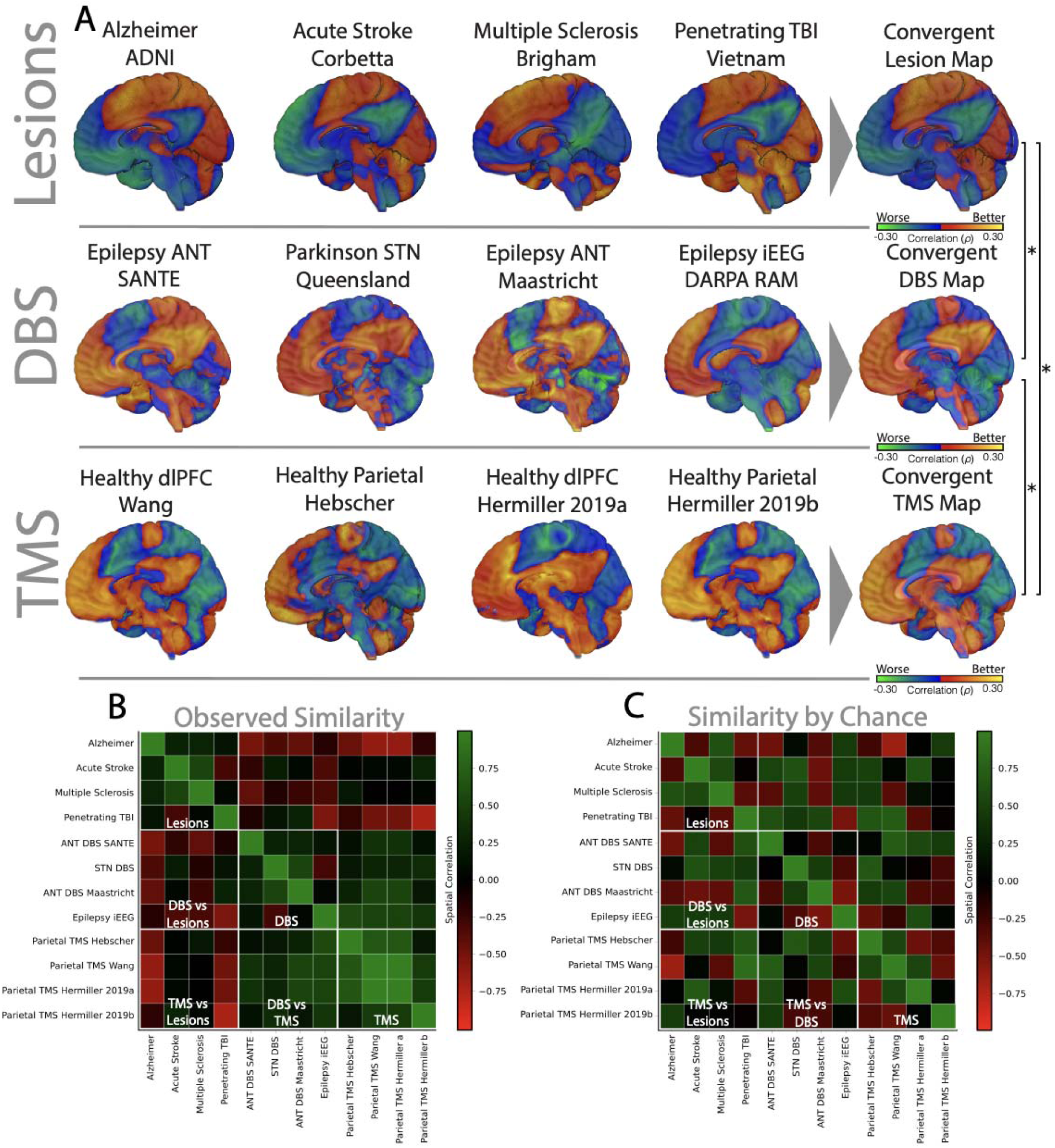
Memory networks from each study converge on similar network distribution. A) Cohort level memory maps (rho-maps) of connections covarying with memory. The rho-maps from DBS, lesions, and DBS were averaged to generate the convergent DBS, lesion, and TMS networks. The convergent network for DBS, lesions, and TMS were more similar than expected by chance, including the similarity between DBS with TMS (Spatial R = 0.80, p = 0.010), DBS with lesions (Spatial R = -0.79, 0.021), and lesions with TMS (Spatial R = -0.68, p = 0.047). The colour scheme for lesional maps were flipped (multiplied by negative one) to emphasize anatomy. B) Spatial correlation of cohort-level memory networks and C) example of chance-level spatial correlations from a single permutation of the cohort-level memory networks. The cohort-level memory networks are on-average more similar than expected by chance (p = 0.034). The cohort-level memory networks were similar within DBS (p = 0.026), lesions (p = 0.047), and TMS (p = 0.006).

We next averaged the cohort-level memory networks within each modality to generate a single lesion-, DBS-, or TMS-based memory network (**Figure 2A, right**). Analyses of these three modality-level networks produced similar results as the analysis of the 12 cohort-level networks: the 3 networks showed similar topography (Absolute Spatial R = 0.75, p = 0.0092), with consistency in sign between the DBS and TMS-based maps (Spatial R = 0.80, p = 0.010), but an inversion in sign between the lesion and DBS-based maps (Spatial R = -0.79, p = 0.021) and between the lesion and TMS-based maps (Spatial R = -0.68, p = 0.047).

Collectively, these results show that lesion locations associated with verbal episodic memory impairment show a similar connectivity pattern as brain stimulation sites associated with verbal episodic memory improvement.

### Deriving a Convergent Memory Network using an Optimized Weighting Algorithm

Next, we examined how best to combine the data from our 12 cohorts to define a convergent memory network. To address this problem, we developed an optimization algorithm to find the weighted average of 11 cohort-level memory networks which explained the most variance within these cohorts and then tested the result in the left-out 12th cohort. This method explained significant variance across the left-out cohorts (U = 115, p = 0.0005), outperformed a simple weighted average of the 11 networks (U = 110.5, p = 0.0276), and explained more variance than connectivity to neuroanatomical structures previously linked to memory such as the hippocampus, circuit of Papez, or default mode network (H = 14.4, p = 0.0252) (Supplementary Figure 2). This was not driven by overfitting due to the optimization algorithm, as the memory network derived from permuted data explained significantly less memory variance than the actual convergent memory network (ΔU = 73.5, p = 0.0001). Interestingly, the method explained more variance in memory outcomes when the memory network included data from all modalities rather than just the specific modality of the left-out dataset (t = 5.1, p = 0.014). The value of including cross-modal data held true when predicting variance in episodic verbal memory outcomes after lesions (t = 6.7, p = 0.011), DBS (t = 5.2, p = 0.018), or TMS (t = 3.2, p = 0.043) (Supplementary Figure 3). After completing the above validation, we then ran this optimized weighting algorithm on the full 12 cohorts to derive a final convergent memory network (**Figure 3A**). To identify network connections that were specific to memory (**Figure 3B**), we performed a permutation analysis and compared results to 21 other non-memory cognitive maps from the same cohorts (Supplementary Figure 4). Results of this analysis were unchanged when controlling for sample size, disease status, modality status, or demographic factors such as age (Supplementary Figure 5).

**Figure 3.**
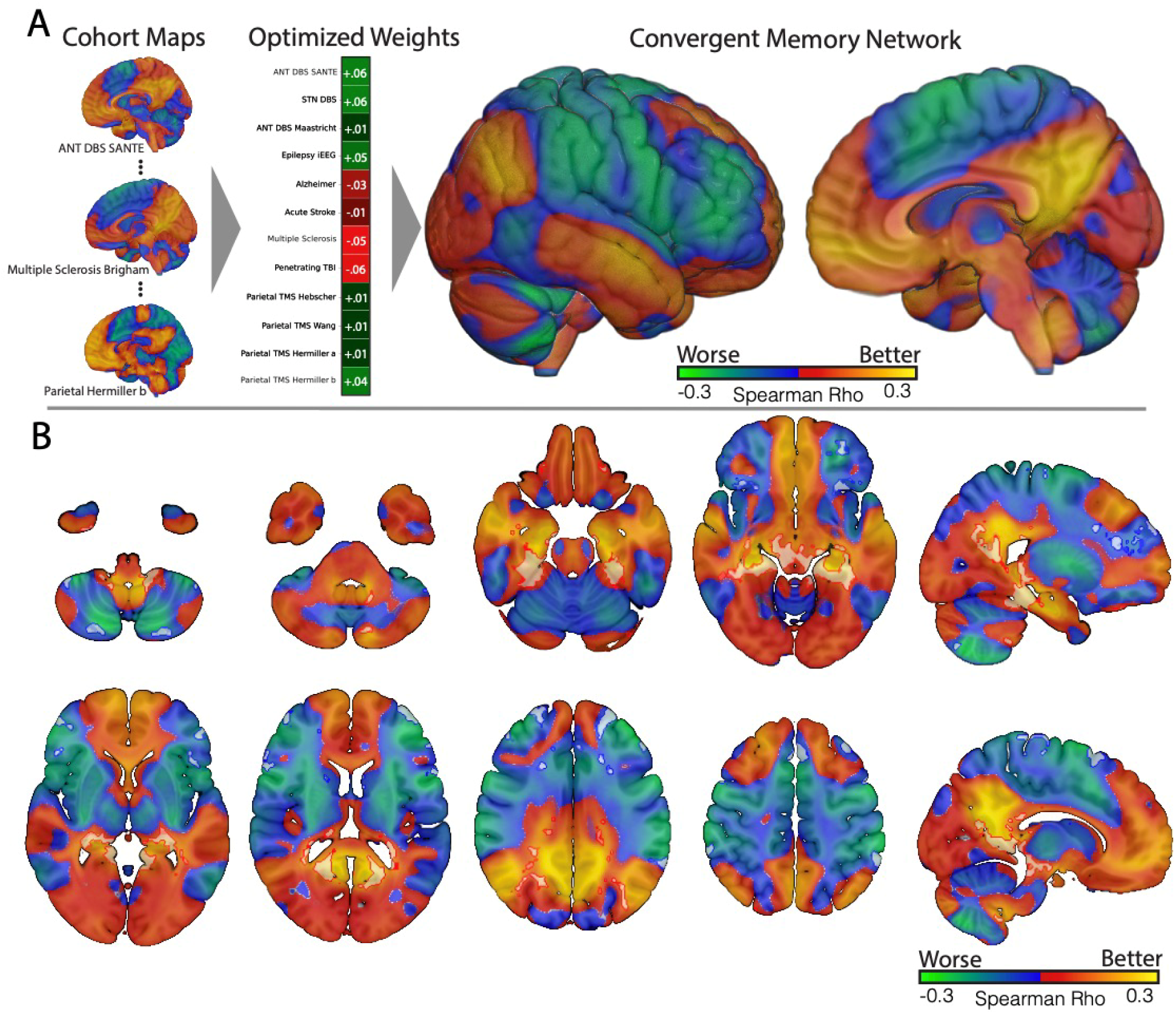
The convergent memory network. A) Derivation of the convergent memory network. The group-level memory maps were averaged with weighting according to their sample sizes, resulting in the whole-brain convergent memory network. B) Mosaic view of the convergent memory network with core nodes overlaid in white. Core nodes were derived from the conjunction of sensitive (consistently strong) with the specific (unique to memory compared with other cognitive domains) correlations across all groups. White regions with outlines represent FWE-significant corrected p-values derived with permutation. The convergent memory network includes core nodes in the cerebellar tonsils, lateral cerebellar hemispheres, hippocampus, retrosplenial cortex, precuneus, lateral parietal cortex, and prefrontal cortex.

### Validating the Convergent Memory Network in Unseen Datasets

To determine if connectivity to the convergent memory network related to memory outcomes in additional independent datasets, we analyzed data from 3 cohorts that were not yet available to us at the time we performed our discovery analysis. These 3 datasets included 31 patients with focal hypometabolism associated with acquired epilepsy (**Figure 4A**), 14 patients with forniceal DBS for Alzheimer Disease (**Figure 4E**), and 16 patients with parietal TMS for non-Alzheimer age-related memory loss (**Figure 4I**), all of whom underwent dedicated episodic verbal memory testing. In these patients, increased connectivity to our convergent memory network was associated worse overall memory scores in patients with hypometabolism (Rho = -0.51, p = 0.0032) (**Figure 4B**), increased memory performance in patients with DBS (Rho = 0.63, p = 0.0124) (**Figure 4F**), and increased memory performance in patients with TMS (Rho = 0.54, p =0.0396) (**Figure 4J**). Compared to their respective modality-specific networks, the convergent memory network explained, on average, 19% more memory variance in hypometabolic sites (t = 12.2, p < 0.0001) (**Figure 4C, D**), 14% more in DBS (t = 14.9, p < 0.0001) (**Figure 4G, H**), and 34% more in TMS (t = 11.9, p < 0.0001) (**Figure 4K, L**). Connectivity to the convergent memory network also explained more variance than connectivity to other neuroanatomical structures previously implicated in memory (Supplementary Figure 6). Results were robust to choice of memory task (Supplementary Table 2).

**Figure 4.**
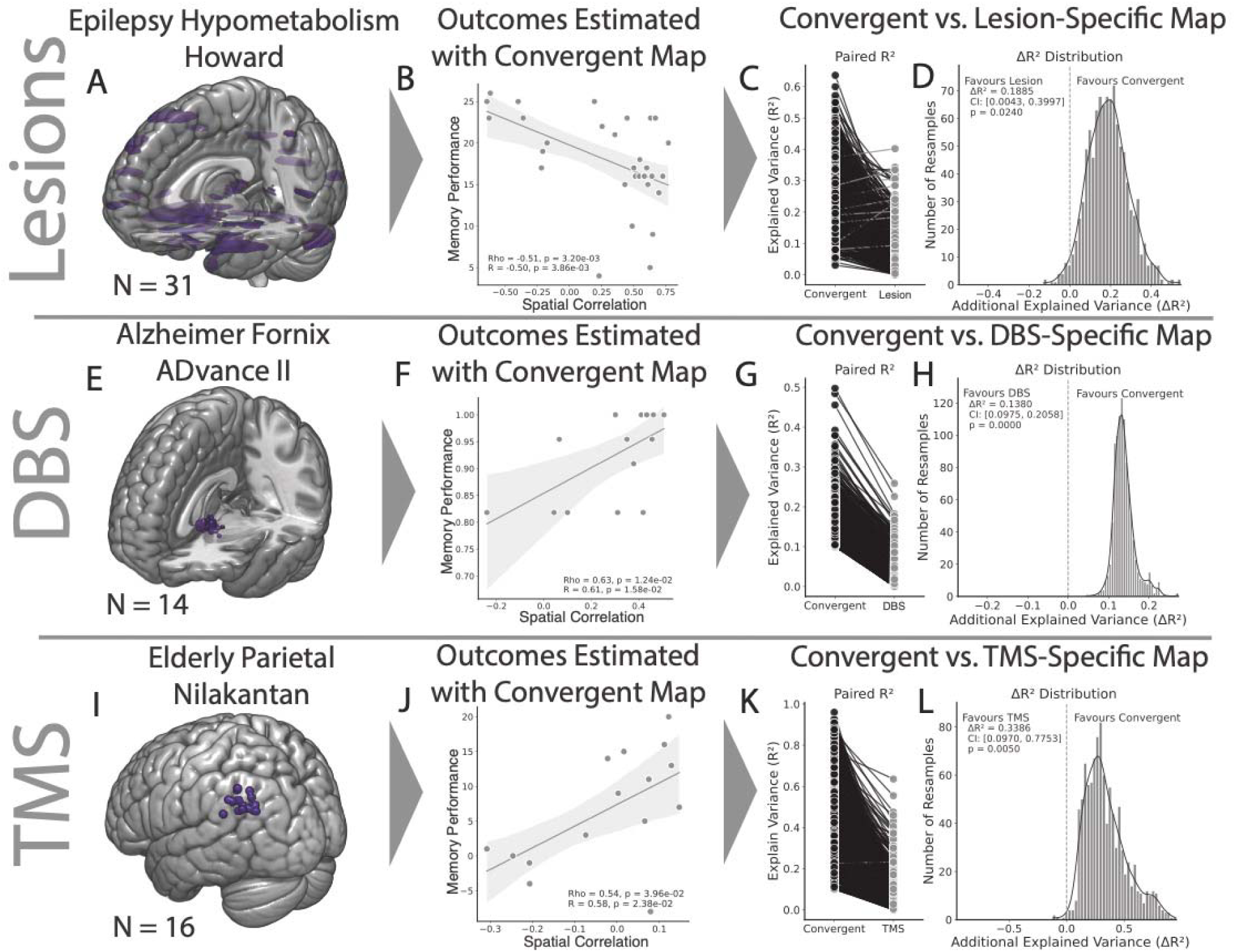
The convergent memory map estimates outcomes better than modality-specific maps. A) The location of focal hypometabolism in 31 patients with epilepsy. B) Similarity of the hypometabolism’s connectivity profile with the convergent memory network was negatively correlated with memory performance (Rho = -0.51, p = 0.0032). C) The convergent memory map explained more memory score variance than the lesion-specific map in 98% of bootstraps. D) The convergent memory map explained significantly more memory variance than the lesion-specific map (mean ΔR^2^ = 0.19, p = 0.0240). E) The location of DBS sites in 14 patients with Fornix DBS for AD. F) Similarity of the DBS site’s connectivity profile with the convergent memory network was positively correlated with memory performance (Rho = 0.63, p = 0.0124). G) The convergent memory map explained more memory variance than the DBS-specific map in 100% of bootstraps. H) The convergent map explained significantly more memory variance than the DBS-specific map (mean ΔR^2^ = 0.14, p = 0.0000). I) The location of TMS sites in 16 elderly patients with age-related memory impairment. J) Similarity of the TMS site’s connectivity profile with the convergent memory network was positively correlated with memory performance (Rho = 0.56, p = 0.0396). K) The convergent memory map explained more memory score variance than the TMS-specific map in 99% of bootstraps. L) The convergent map explained significantly more memory variance than the TMS-specific map (mean ΔR^2^ = 0.34, p = 0.0050). Black bars in paired plots represent bootstraps where the convergent map outperformed modality-specific maps.

### Statistics on the Convergent Memory Network and Optimized Weights

The weights generated by the optimization algorithm were not treated as a free or repeated statistical choice during inference. Instead, cohort weights and their associated signs were determined during model construction within each training fold and were held fixed for all subsequent analyses. The signs were not allowed to vary during testing nor conditionally flipped. Group-level statistical inference was performed exclusively on left-out cohorts using these fixed weights, were not tested multiple times, and no selection was made based on any individual cohort. Voxelwise statistics were subject to multiple comparisons correction using the Westfall-Young procedure while holding weights constant.

### Relation to outcomes of prior brain stimulation trials Alzheimer Disease

To investigate whether our network might help explain heterogeneity in prior brain stimulation trials for Alzheimer Disease, we performed a systematic review of prior DBS and TMS randomized controlled trials for Alzheimer Disease (Supplementary Figure 7). We identified 21 studies which met our inclusion criteria (DBS = 2, TMS = 19, Supplementary Table 3). For each study, we calculated the effect size upon overall cognition (Cohen’s D) and reconstructed the location of the stimulation site (see methods). We then overlaid each stimulation site onto the “memory target network”, a map of voxelwise connectivity to the convergent memory network (**Figure 5A**). Despite targeting at least 7 different brain regions, we found that the majority of studies (16/21) stimulated sites falling within positive regions of the memory target network. The 5 studies that fell outside this target network (i.e. in negative regions) had lower cognitive effect sizes compared to 16 studies that stimulated sites within our network (t = 2.64, p = 0.037) (**Figure 5B**), controlling for covariates such as age, stimulation site, stimulation type, and stimulation frequency. This difference was significant when tested nonparametrically (U = 13, p = 0.025), with outliers removed (t = 2.40, p = 0.025), with DBS studies removed (t = 2.65, p = 0.023), or when restricted to TMS studies targeting the frontal cortex (t = 2.85, p = 0.020).

**Figure 5.**
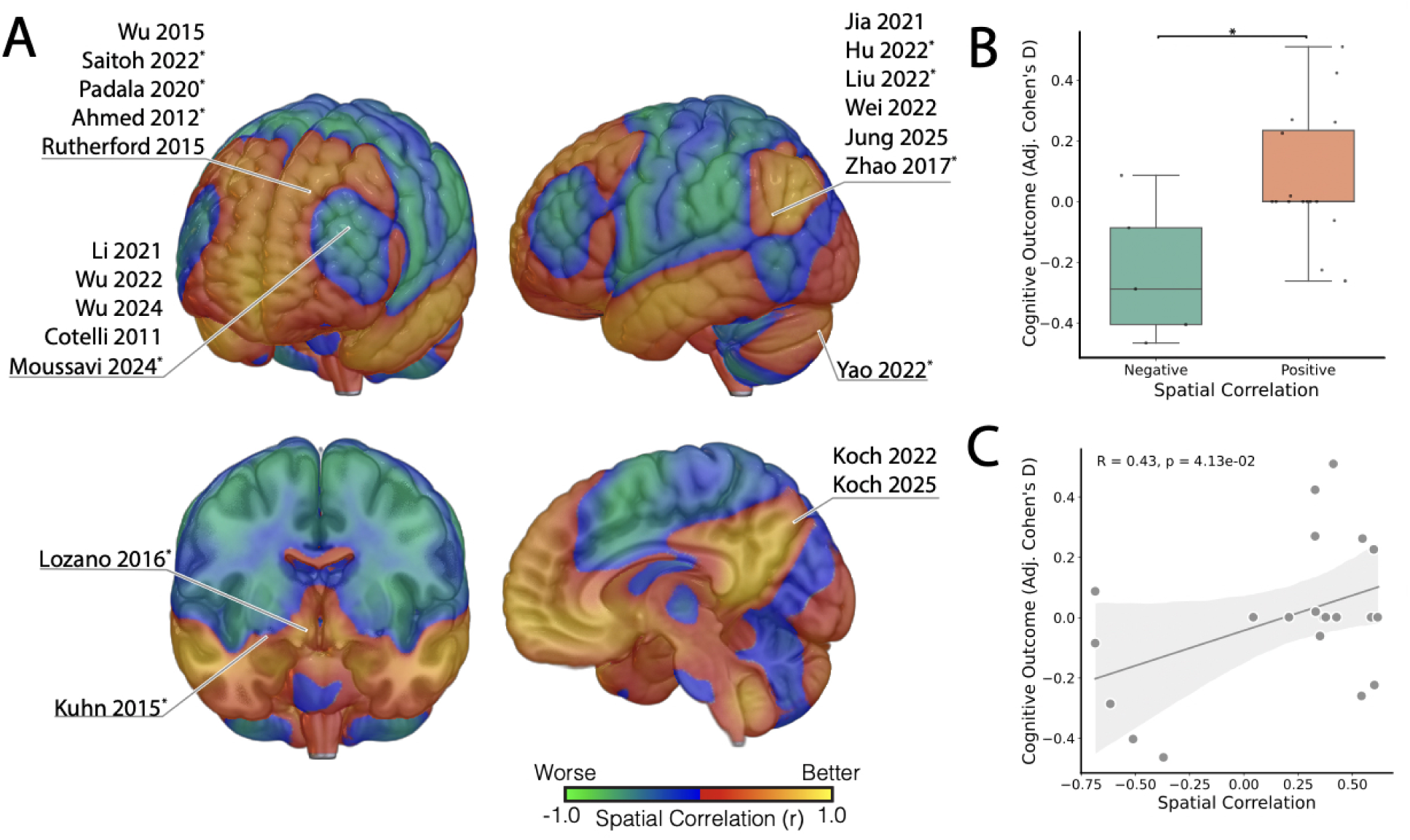
Meta-Analysis Relating Prior Alzheimer Disease Brain Stimulation Studies Outcomes to Memory Target Network. A) Brain regions targeted by 21 stimulation sites from prior randomized-controlled trials of TMS and DBS for Alzheimer Disease. Each stimulation site within a region varied, but were aggregated into a single brain region for visualization. Overlaid on the brain is the memory target network, which represents the connectivity of each voxel to the convergent memory network itself. The memory target network’s most positive regions represent theoretically ideal stimulation targets for memory. Studies annotated with an asterisk (*) had bilateral symmetric stimulation sites. B) Boxplots comparing cognitive outcomes a stimulation sites with negative versus positive spatial correlations with the convergent memory network (green versus yellow regions in panel A) after controlling for covariates. Stimulation sites in positive regions of the memory target network had higher cognitive effect sizes when compared parametrically (t = 2.76, p = 0.012) or nonparametrically (U = 55, p = 0.0013). C) Scatterplot relating each stimulation site’s spatial correlation to the convergent memory network to each study’s cognitive effect size after controlling for covariates. Increasing spatial correlation, or having a stimulation site within the positive regions of the memory target network, was correlated with cognitive outcome (r = 0.43, p = 0.041).

Across all 21 studies, connectivity to the convergent memory network also correlated with cognitive effect size (r = 0.43, p = 0.041) (**Figure 5C**). This analysis remained significant with outliers removed (r = 0.42, p = 0.0072), when redone with permutation (r = 0.43, p = 0.0257), with DBS studies removed (r = 0.44, p = 0.048), or when restricted to TMS studies targeting the frontal cortex (r = 0.64, p = 0.047) (Supplementary Figure 8). Convergent memory network connectivity explained more outcome variance than overlap with an Alzheimer Disease response network derived from DBS patients (Supplementary Figure 9) or other neuroanatomy traditionally associated with memory (Supplementary Figure 10).^32^

## Discussion

Our results suggest that lesions, DBS, and TMS sites influencing verbal episodic memory are connected to a convergent neuroanatomical network. Connectivity to this convergent memory network explained more variance in memory outcomes than connectivity to modality-specific networks or to other structures implicated in memory. Additionally, connectivity of prior Alzheimer Disease DBS and TMS stimulation sites to this convergent memory network correlated with cognitive effect size.

### A Convergent Memory Network Across Modalities

We found that connections with lesion locations, DBS sites, and TMS sites covarying with verbal episodic memory were similar across 15 cohorts spanning 3 modalities. Our finding that memory effects depend on network connectivity is consistent with prior lesion,^29^ DBS,^45^ and TMS^16^ studies of memory. Our study expands this prior work by showing that the connections that covary with memory are the same across these three independent modalities. This convergence is remarkable given large differences in the neuroanatomical location of lesions and brain stimulation sites across cohorts, and the different mechanisms thought to underlie lesion, DBS, and TMS effects on memory.^46,47^ Further, we found that our convergent memory network explained significantly more variance in memory outcomes than modality-specific networks. While prior work has evaluated within-modality and multimodal-networks,^38^ prior to this study, one might have assumed that lesion data would best predict lesion effects, DBS data would best predict DBS effects, and TMS data would best predict TMS effects.

There are a few potential explanations for why multimodal data explained more variance in verbal episodic memory scores than matched-modal data in this study. First, multimodal data may average out confounds inherent in analyses focused on a single modality,^48,49^ leading to more accurate neuroanatomical localization.^26^ Second, the use of multimodal data allowed for more datasets to be included, and more data alone may lead to more accurate localization.^50^ Third, the optimization algorithm we used combined datasets based on explained variance, which may better capitalize on multimodal data compared to prior methods based on weighted averages.^38^ Finally, there was significant heterogeneity across datasets within a modality which may have biased us against finding an advantage of within-modality analyses. Future work is needed to determine the benefit of using multimodal data, and whether the use of the optimization algorithm introduced here extends to other symptoms and disorders.

### Neuroanatomy of the convergent memory network

Many brain regions in our convergent memory network have been consistently implicated in memory, including the hippocampus, parahippocampal gyrus, precuneus, entorhinal, parietal, and prefrontal cortices.^8,16,28,33,51,52^ Other brain regions in our convergent memory network such as the lateral temporal cortex and cerebellum are not part of traditional memory circuits such as the circuit of Papez, but may play important roles in memory. For example, stimulation of the lateral temporal cortex can cause memory flashbacks^53^ and improve memory task performance in traumatic brain injury^54^ while stimulation of the cerebellum can modulate hippocampal activity,^55–57^ and improve memory task performance in Alzheimer Disease.^58^ Beyond individual brain regions, our results support the hypothesis that memory depends of a distributed network of interacting brain regions^29,59–61^ and provides a data-driven network topography for the location of these regions.

### Directionality of effects on memory

Despite similar network topography across our 15 cohorts, the sign of the association varied depending on the modality. Lesions with more connectivity to the convergent memory network were associated with worse memory scores, consistent with prior lesion-based studies.^29,61,62^ In contrast, both TMS and DBS sites with more connectivity to the network were associated with improved memory. The inversion in sign between lesion and TMS datasets is not surprising, given prior work combining these types of datasets^38,63^ and hypotheses that TMS may strengthen network-level functions through neuroplastic effects.^16,47^ However, the inversion in sign between lesion and DBS datasets is unexpected, and not consistent with current mechanistic models of DBS acting as an informational ‘lesion’.^46^ Two potential explanations for the sign inversion between lesion and DBS datasets are: (1) that lesional effect of DBS depends upon the state of the brain, or (2) that the view of DBS as a ‘lesion’ may potentially be oversimplified.

Recent research has suggested the lesional effect of DBS may depend upon the baseline state of the brain. For example, hippocampal stimulation impaired memory in epilepsy patients with intact baseline memory, whereas patients with impaired baseline memory actually benefitted from stimulation.^64^ Similarly, DBS in both Parkinson’s and Alzheimer Disease can impair cognition when the hippocampus is intact, but improve cognition when the hippocampus is atrophic.^65^ It is possible that there is a difference in the rate of baseline memory dysfunction in our lesion versus DBS cohorts based the prevalence of memory dysfunction in these conditions.^66–71^ Unfortunately baseline memory scores were only available for one of our 10 lesion and DBS datasets, so we were unable to test this hypothesis.

Several lines of research have suggested it is possible DBS is not acting like a ‘lesion’ with respect to memory outcomes in our datasets and may instead be inducing more complicated physiological effects. This hypothesis is supported by observations of increased metabolic activity in memory-related regions^20^ and increased hippocampal acetylcholine after fornix DBS,^72^ increased cerebellar grey matter after STN DBS,^73^ and increased long-term potentiation as well as hippocampal metabolism following hypothalamic DBS.^74^ These findings and others suggest that DBS may support rather than inhibit cellular function.^20,45,57,73–76^ Integrating our current results with this prior research, it is possible DBS may be more similar to TMS than to lesions with respect to its influence upon memory anatomy.^47^

### Implications for Clinical TMS

TMS has been used to target at least 10 different brain regions for treatment of verbal episodic memory impairment in Alzheimer Disease, 7 of which were randomized trials and met inclusion criteria in our study.^8–19^ Of these targets, the dorsolateral prefrontal cortex has been targeted most frequently, and was used in the largest trial of TMS for Alzheimer Disease to date.^10,19,77,77–85^ Our study provides potential new insight into the heterogenous outcomes across these trials. Specifically, the TMS sites associated with therapeutic benefit were selected using scalp measurements which may have fallen within more dorsal^86^ positive nodes of the memory target network.^19,80–82^ Conversely, TMS sites selected using neuroimaging were more ventral,^86^ falling within negative regions of the convergent memory network, and were associated with weaker effect sizes.^19,80–82^ Our convergent memory circuit also includes strongly positive nodes in the precuneus and posterior parietal cortex, both of which have recently been successful TMS targets for improving memory and Alzheimer Disease.^8,16,87–90^ Beyond providing insight into the prior TMS literature, our convergent memory circuit may provide a path for optimizing and individualizing TMS targets. Prior work has individualized TMS targets for memory using hippocampal connectivity.^16,47,87,88,91^ However targets based distributed network connectivity rather than a single region have been found to be more robust and reliable.^92^ The convergent memory network presented here may be a network-level target for TMS individualization in future TMS trials.

### Implications for Clinical DBS

It remains unclear how to program DBS electrodes for treatment of memory dysfunction in Alzheimer Disease. Standard DBS programming with the monopolar review may be challenging due to the time-intensive assessments and moment-to-moment variability in human memory.^93^ This raises the possibility of using neuroimaging-based programming in DBS for Alzheimer Disease by maximizing stimulation volume overlap with the memory target network. Although functional connectivity maps have been used to reprogram DBS electrodes in Parkinson Disease to avoid cognitive decline,^45^ the hypothesis that this approach can actually improve cognition in Alzheimer Disease remains to be tested.

The subcortical peaks of the memory target network included the Basal Nucleus of Meynert and fornix, with the posterior parahippocampal gyrus being the whole-brain peak connection. While the Basal nucleus of Meynert and fornix have demonstrated potential in prior trials of Alzheimer Disease DBS,^7,24^ posterior parahippocampal gyrus DBS has not been attempted for Alzheimer Disease. Interestingly, in patients with epilepsy this region been associated with improved associative memory after theta-burst microstimulation,^34^ improved visual memory after right but not left white matter macrostimulation,^94^ and improved verbal as well as visuospatial memory after non-entorhinal parahippocampal grey matter macrostimulation.^95^ This memory improvement is unique among other mesial temporal stimulation sites.^35,95–98^ While it does not escape our notice that this may be a potential Alzheimer Disease DBS target, there are no trials to support implanting DBS based on functional connectivity alone, and given the limited retrospective nature of our results, are not sufficient evidence in themselves to support DBS of the posterior parahippocampal gyrus.

### Limitations

A limitation of note is that this study is retrospective in nature. While we do work with causal sources of information such a lesions or brain stimulation sites influencing memory, retrospective data should still be interpreted with care. This study includes multiple sources of heterogeneity, as many of the cohorts included in this study were not controlled trials, used multiple different cognitive examinations, and were included multiple different diseases.^99^

Additionally, cognition and memory are complex phenomena which are moderated by numerous variables spanning from sleep quality to integrity of the memory network.^41,45,100^ We do not have access to all of these variables in our underlying cohorts, and are therefore unable to evaluate how these influence the localization of the convergent memory network or memory outcomes.

Our analyses are primarily conducted in standardized brain space, which carries the inherent risk of registration inaccuracies in the location of lesions, DBS electrodes, or TMS sites to template brain space. For our 15 datasets, we attempt to mitigate these risks by having lesions manually co-registered by experts; DBS electrodes corrected for brain shift,^101^ were multispectral normalized,^102^ phantom electrode localization validated,^103^ with associated manual refinement of atrophy with warp fields;^104^ and TMS sites were localized directly from reported stimulation coordinates. The risk of inaccuracy in the location of brain stimulation sites is higher for our meta-analysis of prior TMS trials for Alzheimer Disease, as we did not have the raw data from these trials and thus the stimulation sites had to be estimated from the published papers. While some studies reported exact coordinates, others reported targeting approaches based on scalp measurements, the coordinates of which had to then be estimated.

Our findings regarding the signs of lesions, DBS, and TMS may be limited in their generalizability, particularly with respect to other diseases. Prior work has found that the networks associated with depression after both lesions and DBS share similar signs, while TMS has an opposite sign to both.^105^ Future investigations are needed to understand how the effect of lesions, DBS, and TMS vary as a function of disease.

Additionally, we work primarily using normative functional connectivity. This has two associated limitations: normative connectomics and the restriction to functional connectivity. Normative connectomics are based upon a large cohort of healthy patients, not diseased patients nor individualized connectivity profiles.^106^ This analysis has yielded robust findings in prior network mapping studies,^6,22,32,45,107^ and prior work suggests that disease- or patient-specific connectomes result in similar findings as normative connectomes.^6,75,108^ Additionally, we are limited in our ability to derive ‘convergent’ tracts which may mediate memory, as the methods required to define how numerous tracts articulate across the brain have not been developed.

## Online Methods

### Ethics Statement

All procedures conformed to the ethical standards of the Brigham and Women’s Hospital / Harvard Medical School Institutional Review Board. As every dataset had been collected previously for other independent studies, this secondary analysis qualified for exemption from additional informed consent.

### Discovery Cohort

The discovery (training) set comprised 1247 individuals whose brains were perturbed by lesions, deep-brain stimulation (DBS), or transcranial magnetic stimulation (TMS). Of the 984 lesion cases, 376 showed Alzheimer-related atrophy,^41^ 87 had acute ischemic stroke, 426 had multiple-sclerosis lesions,^61,109^ and 96 had penetrating traumatic injuries sustained during the Vietnam War. The 180 DBS cases included: two epilepsy cohorts with electrodes in the anterior thalamic nucleus (n = 68 and n = 16), a Parkinson cohort with subthalamic stimulation (n = 59), and an epilepsy cohort in which stimulation was delivered through multiple intracranial EEG contacts (n = 37). Finally, 82 participants underwent left-parietal TMS across four independent studies (n = 20, 15, 16, and 31). Every study provided high-resolution MRI or CT for precise localization of the lesion or stimulation site and supplied objective scores from validated verbal episodic memory tests. Subjects missing either imaging or behavioral data were excluded. Although no power calculation was performed, this sample is 30 times larger than prior reports which linked cognitive outcomes after lesions^29^ or DBS^45^ to brain networks, and no comparable prior study of retrospective normative TMS memory network mapping.

### Test Cohort

93 patients across 3 test cohorts were collected from collaborators after the convergent memory network had been identified. This data consisted of 31 patients with refractory epilepsy who exhibited focal hypometabolism on PET (45±12 years old) and memory sub-scores from the Autonomous Cognitive Examination;^110^ 14 patients with Alzheimer disease who received fornix DBS (68±8 years old) and delayed memory scores from the 11-item Alzheimer Disease Assessment Scale–Cognitive Subscale (ADAS-Cog 11) as well as memory subscales;^20^ and 16 older adults with non-Alzheimer age-related memory complaints who received parietal TMS (73±4 years old) and a delayed-recall battery.^91^ Patients without a delayed recall test, or without a localizable lesion or stimulation site, were excluded.

### Evaluating if Memory Outcomes Can be Explained Without Networks

We first investigated if memory outcomes could be explained by local anatomy like the hippocampus or Circuit of Papez. Within each cohort, we performed a traditional voxel-lesion symptom map, where we correlated the involvement of each voxel to memory outcomes. We measured the overlap of each patients’ lesion or stimulation sites with structures such as the hippocampus or Circuit of Papez and correlated these with memory outcomes in each cohort. We then measured how similar these voxel-lesion symptom maps were across cohorts, and evaluated if involvement of these maps could cross-predict memory outcomes across cohorts. We also evaluated if overlap of lesions or stimulations with anatomy typically associated with memory, such as the hippocampus or Circuit of Papez, correlated with memory outcomes.

### Deriving Cohort-Level Memory Maps

We implement network mapping, an approach which has found success in localizing distributed neuropsychiatric symptoms to brain networks as opposed to single anatomical locations.^111^ For every participant, we overlaid the lesion mask or stimulation volume onto a normative resting state functional-connectivity atlas derived from 1,000 healthy volunteers (Brain Genomics Superstruct Project).^43,44^ The procedure generated a whole-brain map that quantified, voxel by voxel, how strongly the focal site was functionally connected to every other brain region. To generate memory maps within each cohort, we performed a voxelwise Spearman correlation relating connectivity strength and verbal memory performance. This process yielded a voxelwise map of the association between connectivity and memory performance, which we call a ‘memory map’. This process was repeated for each of the 12 cohorts, generating a cohort-level ‘memory map’ per dataset.

### Similarity among Cohort-Level Memory Maps within a modality

To test whether the 12 cohort-level memory maps were converging upon similar networks, we computed the mean spatial correlation between every pair of maps and compared that value against a null distribution composed by 10,000 permutations of the same process. We repeated the procedure separately for lesion-to-lesion, DBS-to-DBS, TMS-to-TMS, and cross-modality comparisons. Lastly, we created an average of the cohort-level memory maps for each modality, one for lesions, one for DBS, and one for TMS. We measured the pairwise similarity across these networks and evaluated significance using a permutation test.

### Generating the Convergent Memory Network

As it is unclear how to converge the 12 cohort-level memory maps into a single network, we developed an optimization algorithm which finds the weighted average best explained memory variance across the 12 underlying cohorts. The algorithm first quantified how strongly each subject’s connectivity profile matched a candidate weighted network (Supplementary Equation 1), then related these similarity values to memory outcomes within each cohort (Supplementary Equation 2). The cohort weights were optimized to produce the weighted average map that maximized explained variance across all cohorts (Supplementary Equation 3). Note that a cohort-level map could be weighted negatively, allowing for sign inversion between maps derived from lesion cohorts versus the TMS and DBS cohorts. To prevent overfitting, the optimized weights were then blended with an unweighted average (Supplementary Equation 4). Optimization was performed using adaptive moment gradient ascent^112^ with numerical differentiation and no assumption of convexity. The final weights were then used to average the 12 cohort-level memory maps into a single convergent network. The 12 cohorts served exclusively as the training dataset, and the 3 held-out cohorts were never used during training or algorithm tuning. To evaluate performance of the convergent network within the training cohort, leave-one-cohort-out cross validation was used. The optimization algorithm was run on 11 cohorts, then applied to the left-out cohort. This was repeated for each cohort.

### Identify Peak nodes of the Convergent Memory Network

We next derived the core nodes of the convergent memory network. To do so, we used a whole-brain permutation analysis of linear models to find connections which were: (1) common to memory, and (2) specific to memory. To identify connections commonly related to memory, we performed a voxelwise 1-sample t-test of the cohort-level memory maps, using the weights derived from the above optimization algorithm. To identify connections specific to memory, we performed a voxelwise 2-sample t-test comparing the 12 cohort-level memory maps (with the above weights) with 21 other non-memory cognitive maps derived from the same cohorts (holding the weights constant for each cohort). These non-memory cognitive maps included visuospatial, language, and executive function. We then performed a conjunction analysis to identify the connections which were both common and specific for memory function across the 12 cohorts. The permutation analysis of linear models utilized a weighted regression, using weights from the above optimization algorithm.

### Testing in Unseen Patients

For every subject in the 3 held-out cohorts, we seeded each individual’s lesion or stimulation site to derive whole-brain functional connectivity. We then measured the spatial correlation between that connectivity profile and the convergent memory network. This yielded a similarity score, which we correlated to cognitive outcomes in the held-out lesion, DBS, and TMS test cohorts (two-tailed, allowing for either a positive or negative correlation). The test cohorts were not evaluated until the convergent memory network was finalized. We repeated this process, but used the training datasets to derive optimized modality-specific networks built from lesions-, DBS-, or TMS-studies. This generated modality-specific convergent memory networks. We repeated the above correlation analysis using these modality-specific maps. We then repeated the analysis where we evaluated how well spatial correlation to the modality-specific maps correlated with memory outcomes within and across modalities. We compared the convergent and modality-specific maps by bootstrapping this correlation value 1,000 times, and then measured the proportion of times one network explained more memory variance than the other. This analysis was also repeated with a priori maps such as the default mode network and Circuit of Papez.^27,28^

### Meta Analysis of Prior Brain Stimulation Studies for Alzheimer Disease

To identify previous randomized controlled trials reporting the effects of brain stimulation in Alzheimer Disease, we performed a systematic review of the literature. This search was conducted in accordance with Preferred Reporting Items for Systematic Reviews and Meta-Analyses (PRISMA) guidelines. We searched PubMed, PsycINFO, and Scopus databases. Keywords searched for all DBS and TMS studies for Alzheimer Disease (Supplementary Table 1). Inclusion criteria limited studies to high quality randomized controlled trials in Alzheimer Disease and sufficient data to calculate effect sizes and recover stimulation sites (Supplementary Table 2). For all included studies, Cohen’s d was calculated using the post-treatment comparison between experimental and control cohorts. Effect sizes were aligned with prior effect sizes reported in the literature.^113^ When multiple cognitive scales were reported, we prioritized the Alzheimer Disease Assessment Scale, then the Clinical Dementia Rating, the Montreal Cognitive Assessment, and lastly the Mini-Mental Status Examination. The systematic review was performed using Review PyPer, an open-access tool used to facilitate full PRISMA-based systematic reviews assisted by large language models.^114^ All results from the systematic review were validated by authors CWH and EGS.

Stimulation sites were estimated several ways, similar to prior validated methods . For studies with reported stimulation coordinates, these were warped into MNI space if not already there, and then used directly. For studies reporting stimulation at a specific electroencephalography contact, a standard mapping between electroencephalography contact and MNI coordinate was used.^115^ For studies reporting craniometric approaches, such as movement of several centimeters after adjusting stimulation threshold at the hand knob of the motor cortex, these movements were applied directly from the MNI space hand knob to derive the final stimulation location. Studies reporting a stimulation location by visualization of a specific target site within a brain, but with no reported coordinate not craniometric approach, were estimated by identifying the corresponding target within the MNI brain. Stimulation sites were defined as falling within a given brain region when they fell within a 5mm radius of each other.

### Comparing Prior Studies to a Targeting Map for the Convergent Memory Network

To derive a map that could potentially serve as a target for brain stimulation, we computed the spatial correlation between the connectivity of every brain voxel and our convergent memory network, using previously validated methods.^116^ This generates a voxelwise map that identifies brain regions that have the same connectivity profile as lesion or stimulation sites that influence verbal memory. We next measured how much the stimulation sites of previously published studies overlapped the memory target network. We then related these measurements to each study’s cognitive outcome effect size.

### Statistical Analysis and Software

All pipelines were executed in Python 3.10 with the open-source CircuitPyper package, which was developed for this manuscript is and is available at https://github.com/Calvinwhow/CircuitPyPer.git. The package depends on Nilearn 0.10.1 for image manipulation, Statsmodels 0.14.0 for statistical tests, and Scikit-learn 1.3.0 for machine-learning routines. Continuous variables are reported as mean ± standard error. All p values are two-tailed; permutation tests used 10,000 iterations and bootstraps used 1,000 resamples. Because datasets were collected independently and could not be assumed identically distributed, primary statistics were computed within each dataset and only then combined at the group level. Composite statistics, such as checking if several correlations were higher than expected, were calculated using permutation testing. All spatial correlations used Pearson correlations and were two-tailed. Pearson correlations were used to evaluate the relationship between connectivity and outcomes throughout the manuscript. To evaluate if Pearson correlations were driven by outliers, each Pearson correlation was repeated with outliers removed using the interquartile range method.

## Data Availability

All data produced in the present study are available upon reasonable request to the authors.

## Acknowledgements

A.H. was supported by the Schilling Foundation, the German Research Foundation (Deutsche Forschungsgemeinschaft, 424778381 – TRR 295), Deutsches Zentrum für Luft-und Raumfahrt (DynaSti grant within the EU Joint Programme Neurodegenerative Disease Research, JPND), the National Institutes of Health (R01MH130666, 1R01NS127892-01, 2R01 MH113929 & UM1NS132358) as well as the New Venture Fund (FFOR Seed Grant). M.D.F. has intellectual property on the use of brain connectivity imaging to analyze lesions and guide brain stimulation, has consulted for Magnus Medical, Soterix, Abbott, Boston Scientific, Tal Medical, MDC Venture Capital, and is on the Scientific Advisory Board of Salma Health. He has received research support from Neuronetics and Boston Scientific. C.W.H. holds a leadership position in CogNet Inc., and has intellectual property related to guiding brain stimulation with connectomic imaging.

