## Supplementary Materials for "A Network Target for Memory Dysfunction Derived from Brain Lesions and Stimulations"

**Supplementary Table 1. Control analyses for similarity of network topology across 12 cohort-level memory networks.**

| **Memory Network Transform** | **Average Metric** | **P-Value** |
| --- | --- | --- |
| None | Pearson r = 0.43 | p = 0.002 |
| None | Spearman Rho = 0.46 | p = 0.001 |
| None | Cosine Similarity = 0.81 | p = 0.0008 |
| Absolute Value | Pearson r = 0.41 | p = 0.047 |
| Absolute Value | Spearman Rho = 0.49 | p = 0.021 |
| Absolute Value | Cosine Similarity = 0.83 | p = 0.0009 |
| Squared Value | Pearson r = 0.51 | p = 0.013 |
| Squared Value | Spearman Rho = 0.57 | p = 0.008 |
| Squared Value | Cosine Similarity = 0.75 | p = 0.005 |

**Supplementary Table 2. Control analyses for choice of memory task in held-out datasets.**

| **Dataset** | **Memory Test** | **Correlation (Rho)** | **Significance (p-value)** |
| --- | --- | --- | --- |
| Lesion | 3-word delayed recall | -0.31 | 0.015 |
| Lesion | 8-word delayed recall | -0.70 | 0.0005 |
| DBS | Cancellation reminders | 0.42 | 0.041 |
| DBS | Delayed word recall | 0.22 | 0.11 |
| DBS | Semantic recall | 0.49 | 0.033 |
| TMS | N/A | N/A | N/A |

**Supplementary Table 3. Study data for systematic review of TMS and DBS randomized-controlled studies for Alzheimer Disease.** AG: angular gyrus. BNM: basal nucleus of Meynert. dlPFC: dorsolateral prefrontal cortex. Studies reporting multiple experimental cohorts in the same study report each cohort in separate rows. iTBS: intermittent theta burst stimulation.

| **Study** | **Stimulation site** | **Stim Type** | **Diagnostic Criteria** | **Diagnosis** | **Mean Age (Std)** | **CDR** | **N** | **M:F** | **Education Years (Std)** | **Test** | **Cohen's d** |
| --- | --- | --- | --- | --- | --- | --- | --- | --- | --- | --- | --- |
| Koch 2025 | Precuneus | rTMS | IWG | mild-to-moderate | 74 (5.1) | 0.5–1 | 27 | 0.52 | 10.2 (4.4) | ADAS | 0.374 |
| Koch 2022 | Precuneus | rTMS | IWG | mild-to-moderate | 75 (5.6) | 0.5–1 | 25 | 0.56 | 10.2 (4.4) | ADAS | 0.359 |
| Liu 2022 | Bilateral AG | rTMS | NIA-AA | probable | 67.28 (7.74) | 0.5–1 | 25 | 0.48 | 10.44 (3.33) | ADAS | 0.500 |
| Li 2021 | Left dlPFC | rTMS | DSM-5 | any | 65.97 (8.47) | NA | 37 | 0.459 | 5.65 (3.21) | ADAS | 0.030 |
| Ahmed 2012 | Bilateral dlPFC | rTMS | NINCDS-ADRDA | probable | 65.9 (5.9) | NA | 11 | 0.50 | 0.9-10 | MMSE | 0.93 |
| Wei 2022 | Left parietal | rTMS | DSM-5 | probable | 70 (8.68) | 0.5–2 | 29 | 0.69 | 7.34 (5.54) | MMSE | 0.101 |
| Yao 2022 | Bilateral cerebellum | rTMS | NIA-AA | any | 63.87 (6.85) | 0.5-2 | 15 | 0.467 | 10.53 (3.8) | ADAS | 0.703 |
| Hu 2022 | Bilateral AG | rTMS | NIA-AA | probable | 76.86 (6.07) | 2 | 21 | 0.619 | 13.05 (3.67) | ADAS | 0.700 |
| Padala 2020 | Left dlPFC | rTMS | Clinical exam | any | 74.3 (5.7) | NA | 9 | 0.111 | NA | MMSE | 0.280 |
| Cotelli 2011 | Left dlPFC | rTMS | NINCDS-ADRDA | probable moderate | 71.2 (6.1) | NA | 5 | NA | 6.4 (1.3) | MMSE | -0.088 |
| Saitoh 2022 | Bilateral dlPFC | rTMS (120% RMT) | NINCDS-ADRDA | probable | 76.2 | 1–2 | 15 | 0.467 | 13.7 | ADAS | 0.259 |
| Saitoh 2022 | Bilateral dlPFC | rTMS (90% RMT) | NINCDS-ADRDA | probable | 77.2 | NA | 13 | 0.769 | 12.2 | ADAS | 0.179 |
| Wu 2015 | Left dlPFC | rTMS | NINCDS-ADRDA | probable | 71.4 (4.9) | NA | 26 | 0.615 | NA | ADAS | 0.741 |
| Jia 2021 | Left parietal | rTMS | DSM-5 | mild-to-moderate | 71.41 (8.85) | 0.5–2 | 35 | 0.714 | 7.7 (5.26) | MMSE | 0.143 |
| Moussavi 2024 | Bilateral dlPFC | rTMS | NA | any | 73.3 (6.9) | 1–2 | 53 | 0.963 | NA | ADAS | -0.337 |
| Jung 2025 | Left parietal | rTMS | NIA-AA | MCI / probable | 69.8 (9.1) | 0.5–2 | 18 | 0.6 | NA | ADAS | 0.481 |
| Lozano 2016 | Bilateral fornix | DBS | NIA-AA | mild | 69.8 (8.1) | 0.5-1 | 30 | 0.67 | NA | ADAS | 1.039 |
| Kuhn 2015 | Bilateral BNM | DBS | NIA-AA / DSM-IV | mild-to-moderate | 69.5 (7.7) | 0.5–1 | 6 | 0.6 | NA | ADAS | 0.187 |
| Wu 2024 | Left dlPFC | rTMS (iTBS) | NIA-AA | probable | 66.8 (8.84) | 0.5-2 | 20 | 0.43 | 10.25 (3.3) | CDR | 0.648 |
| Wu 2022 | Left dlPFC | rTMS (iTBS) | NINCDS-AD | probable | 66.5 (8.2) | 0-2 | 24 | 0.58 | 9.7 (4.7) | MoCA | 0.47 |
| Rutherford 2015 | Bilateral dlPFC | rTMS | Clinical exam | probable | 57 (7.2) | NA | 10 | NA | NA | ADAS | 0.64 |

**Supplementary Table 4. Search terms for systematic review of TMS and DBS randomized-controlled studies for Alzheimer Disease.**

| **Brain-stimulation search terms** | **Alzheimer-disease search terms** |
| --- | --- |
| transcranial magnetic stimulation | Alzheimer disease |
| TMS | Alzheimer’s disease |
| rTMS | Alzheimer* |
| theta-burst stimulation | dementia of the Alzheimer type |
| TBS | AD dementia |
| deep brain stimulation |  |
| DBS |  |
| intracranial electrical stimulation |  |
| intracranial stimulation |  |
| non-invasive brain stimulation |  |
| NIBS |  |
| invasive brain stimulation |  |
| neuromodulat* |  |
| neurostimulation |  |
| brain stimulation |  |

**Supplementary Table 5. Criteria for systematic review of TMS and DBS randomized-controlled studies for Alzheimer Disease.**

**Criteria**

Human research

Randomized controlled trial

Control data reported

Mild/moderate/severe Alzheimer Disease

Transcranial magnetic stimulation

Deep brain stimulation

Clear stimulation site

Results not duplicated from prior publication

Not inhibitory stimulation

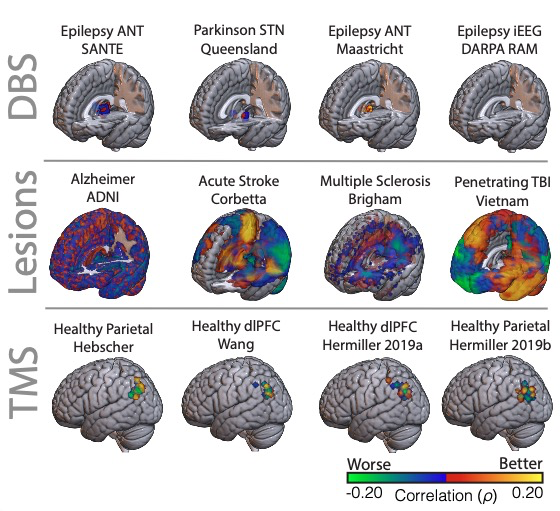
 **Supplementary Figure 1. Voxelwise lesion-symptom maps relating lesion or stimulation site to memory outcomes.** Top) DBS sites influencing memory across 4 datasets, including the anterior nucleus of the thalamus in epilepsy (n = 68, n = 16), the subthalamic nucleus in Parkinson Disease (n = 59), and multifocal iEEG in epilepsy (n = 37). Middle) Lesion sites influencing memory across 4 datasets, including atrophy In Alzheimer Disease (n = 376), acute strokes (n = 87), multiple sclerosis (n = 862), and penetrating traumatic brain injury (n = 96). Bottom) TMS sites influencing memory across 4 datasets, including parietal stimulation in healthy controls (n = 20, n = 15, n = 16, n = 31).

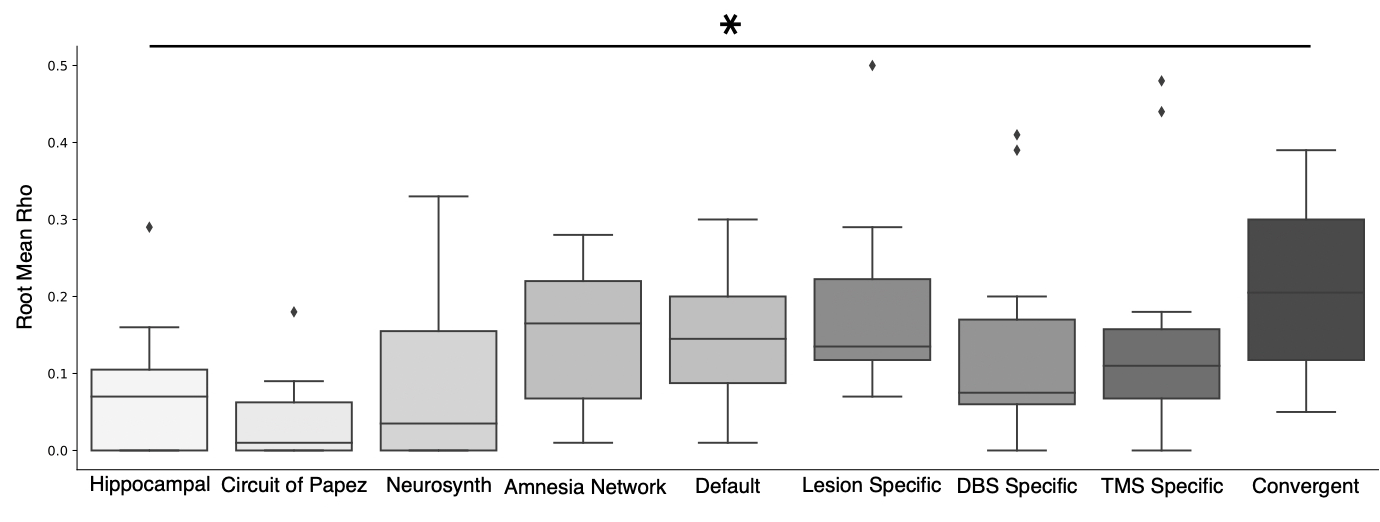

**Supplementary Figure 2. Convergent memory network explains most variance in the leave-one-cohort-out cross-validation.** Each boxplot represents the median and interquartile range of correlation coefficients across all 12 cohorts. The lesion-specific, DBS-specific, TMS-specific, and convergent memory network boxplots were calculated using leave-one-cohort-out cross validation. There was a significant difference across the distributions found by Kruskall-Wallis testing (H = 14.4, p = 0.0252), which was driven by higher performance in the convergent memory network.

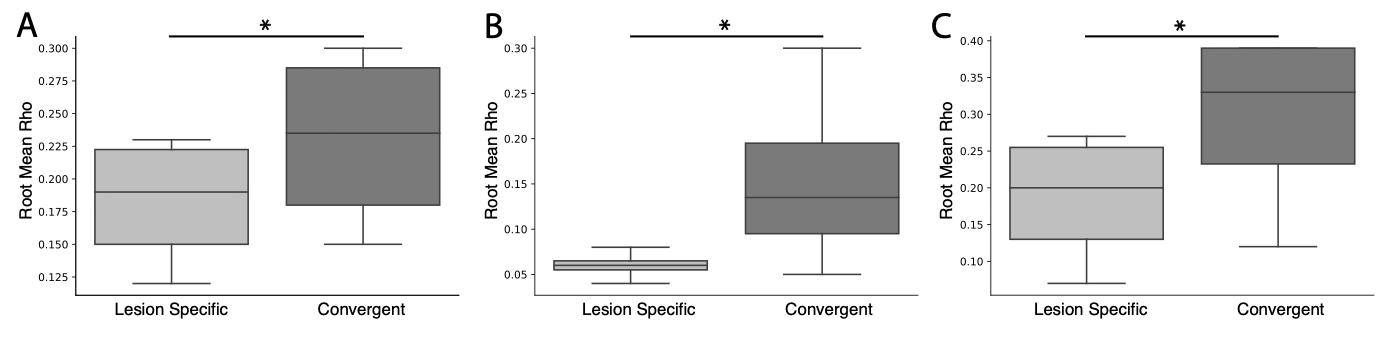
**Supplementary Figure 3. Convergent memory network outperforms modality-specific networks.** Each boxplot represents the median and interquartile range of correlation coefficients across all 12 cohorts. The lesion-specific, DBS-specific, TMS-specific, and convergent memory network boxplots were calculated using leave-one-cohort-out cross validation. A) Comparison between correlations only within the lesion cohorts using the lesion-specific and convergent memory networks. The convergent memory network outperformed the lesion-specific network (t = 6.7, p = 0.011). B) Comparison between correlations only within the DBS cohorts using the DBS-specific and convergent memory networks. The convergent memory network outperformed the DBS-specific network (t = 5.2, p = 0.018). C) Comparison between correlations only within the DBS cohorts using the DBS-specific and convergent memory networks. The convergent memory network outperformed the DBS-specific network (t = 3.2, p = 0.043).

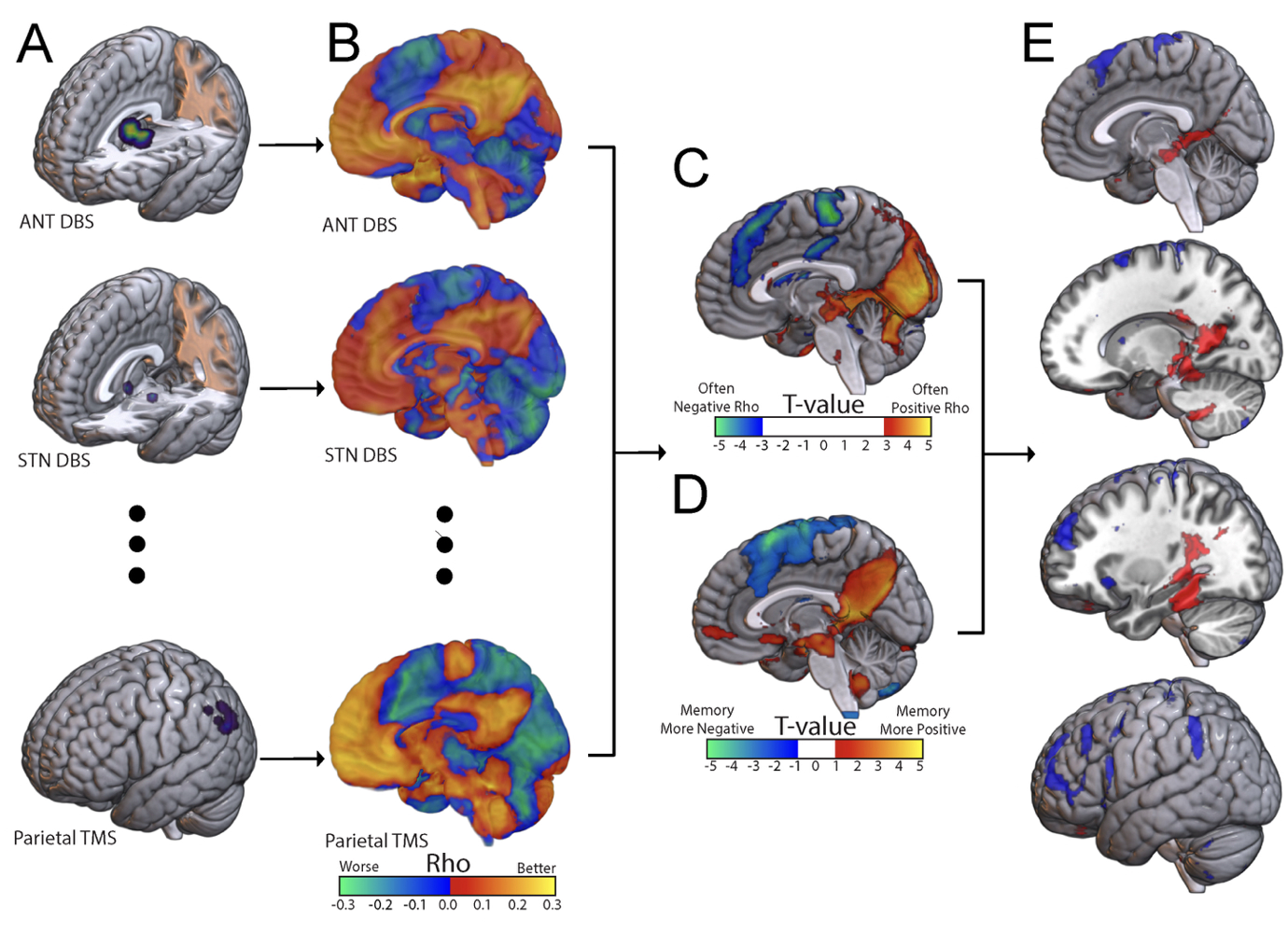
**Supplementary Figure 4. Sensitive and Specific Connections Covarying with Memory.** A) Group-level DBS, lesion, and TMS sites across the 12 discovery cohorts. B) Group-level Rho-maps derived across each cohort. Maps were generated by seeding whole-brain connections covarying with DBS, lesion, and TMS sites. Within each cohort, whole-brain connections were Spearman correlated with memory outcomes, resulting in the Rho Map per cohort. C) Sensitivity map demonstrating group-level Rho Values which were consistently positive or negative across the 12 group-level Rho maps. Lesional Rho Maps were multiplied by negative one prior to evaluation. Maps are thresholded to FWE-corrected significant p-values. D) Specificity map, demonstrating connections which are unique to memory, compared to 21 other cognitive maps derived from the same cohorts (visuospatial, language, and executive function). Lesional Rho Maps were multiplied by negative one prior to evaluation. Maps are thresholded to FWE-corrected significant p-values. E) Conjunction of the FWE-corrected significant p-values in the sensitivity and specificity maps. Nonzero regions have consistently strong connections across all group-level maps while simultaneously being unique compared to other cognitive domain maps.

**Supplementary Figure 5. Network Topology Unchanged After Controlling for Covariates.** Whole-brain voxelwise analysis to identify core nodes of the convergent memory network controlling for covariates such as sample size, disease status, modality status, or demographic factors such as age. A) Group-level DBS, lesion, and TMS sites across the 12 discovery cohorts, along with their B) group-level Rho-maps. C) The covariate-controlled sensitivity map of connections consistently related to memory outcomes across the 12 cohorts was more similar to the uncontrolled sensitivity map than expected by chance (Spatial R = 0.68, p = 0.044). D) The covariate-controlled specificity map was more similar to the uncontrolled specificity map than expected by chance (Spatial R = 0.78, p = 0.0087). E) The covariate-controlled conjunction map was more similar to the uncontrolled conjunction map than expected by chance (Dice coefficient = 0.86, p < 0.0001).

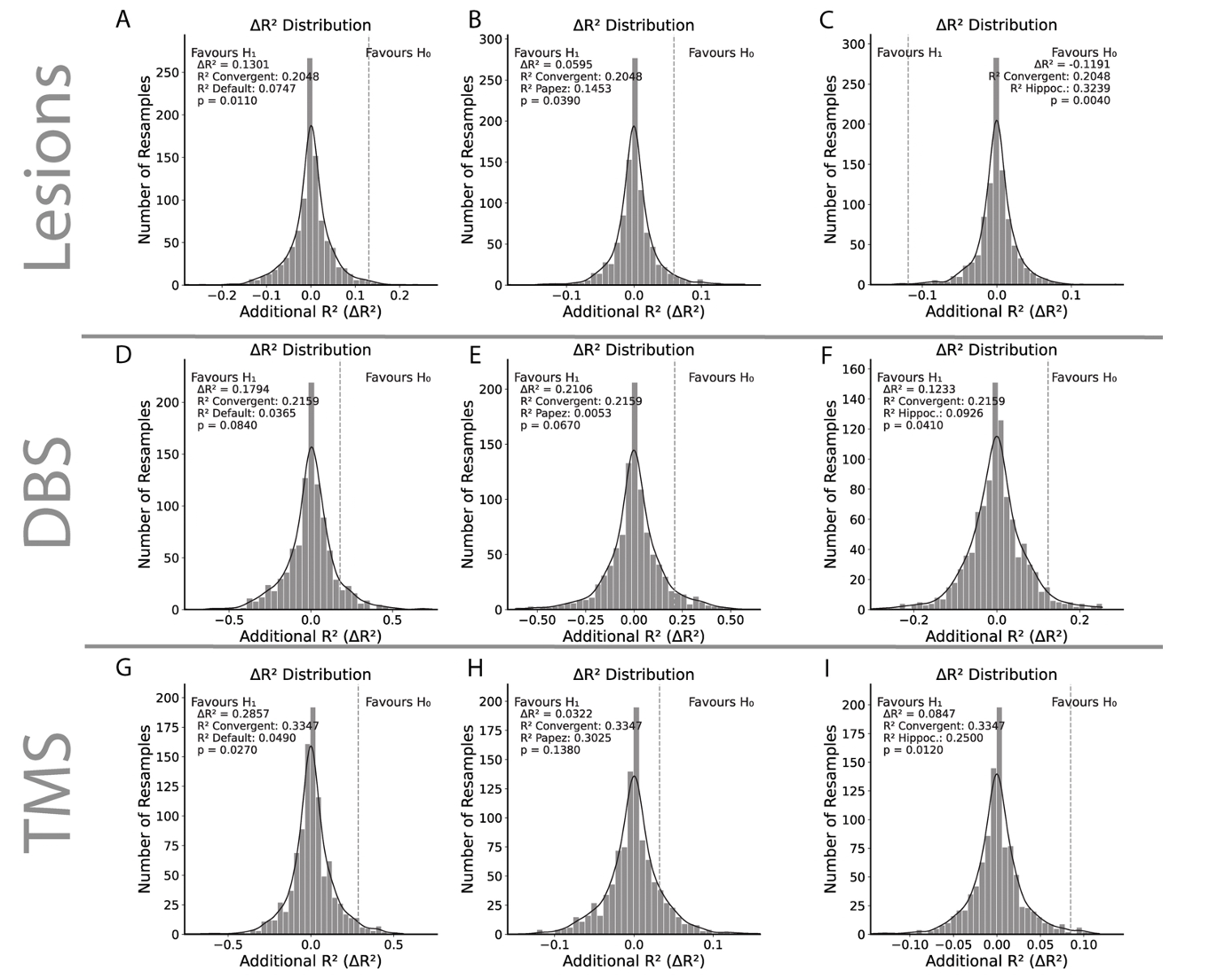
**Supplementary Figure 6. Convergent Memory Network Outperforms Other Memory-Related Neuroanatomy in Relating to Memory Outcomes in Unseen Data.** Distribution of ∆R^2^ across permutation tests, with dashed line representing observed ∆R^2^ across connectivity to the convergent memory network versus other memory-related anatomy. On average, across lesions, DBS, and TMS, the convergent memory network explains more variance than connectivity to the Default Mode Network, Circuit of Papez, or hippocampus (∆R^2^ = 0.11, p = 0.0049). In lesions, connectivity to the convergent memory network explains more variance in memory outcomes than A) the Default Mode Network (∆R^2^ = 0.13, p = 0.011) and B) the Circuit of Papez (∆R^2^ = 0.060, p = 0.039), but not C) the hippocampus (∆R^2^ = -0.12, p = 0.0040). In DBS, connectivity to the convergent memory network explains more variance in memory outcomes than D) the Default Mode Network (∆R^2^ = 0.18, p = 0.0840), E) the Circuit of Papez (∆R^2^ = 0.21, p = 0.067), and F) the hippocampus (∆R^2^ = 0.12, p = 0.041). In lesions, connectivity to the convergent memory network explains significantly more variance in memory outcomes than G) the Default Mode Network (∆R^2^ = 0.29, p = 0.027), H) the Circuit of Papez (∆R^2^ = 0.032, p = 0.14), and I) the hippocampus (∆R^2^ = 0.085, p = 0.012). All p-values for ∆R^2^ analyses were calculated from 1,000 permutations.

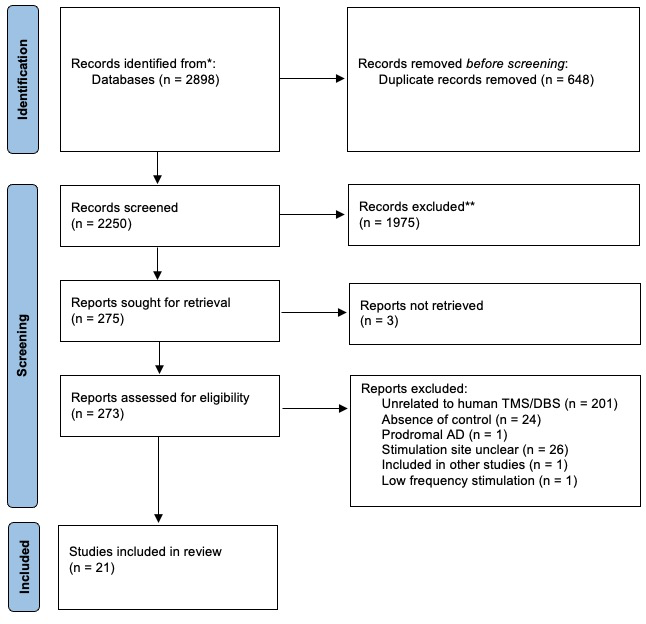
 **Supplementary Figure 7. PRISMA flowchart for systematic review of TMS and DBS randomized-controlled studies for Alzheimer Disease.** Out of 2898 records, 2250 titles and abstracts were screened, 273 full reports were evaluated for inclusion, and 21 studies were ultimately

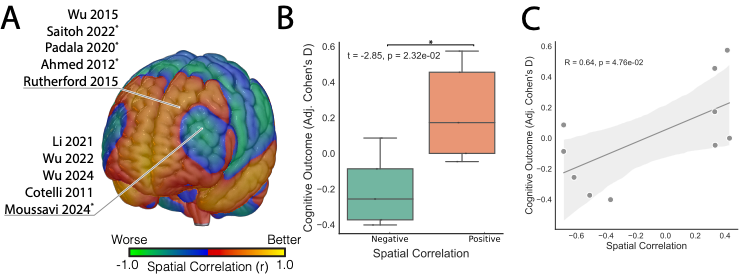
**Supplementary Figure 8. Comparison of Dorsolateral Prefrontal Cortex Brain Stimulation Studies Outcomes Using Memory Target Network.** A) Stimulation locations from 10 prior randomized-controlled trials of TMS for Alzheimer Disease, specifically within the dorsolateral prefrontal cortex. Overlaid on the brain is the memory target network, which represents the connectivity of each voxel to the convergent memory network itself. The memory target network’s most positive regions represent theoretically ideal stimulation targets for memory. Studies annotated with an asterisk (*) had bilateral symmetric stimulation sites. B) Boxplots comparing cognitive outcomes a stimulation sites with negative versus positive spatial correlations with the convergent memory network (green versus yellow regions in panel A) after controlling for covariates. Stimulation sites in positive regions of the memory target network had higher cognitive effect sizes when compared parametrically (t = 2.85, p = 0.029) or nonparametrically (U = 2.00, p = 0.032). C) Scatterplot relating each stimulation site’s spatial correlation to the convergent memory network to each study’s cognitive effect size after controlling for covariates. Spatial correlation correlated with cognitive outcomes (r = 0.64, p = 0.048).

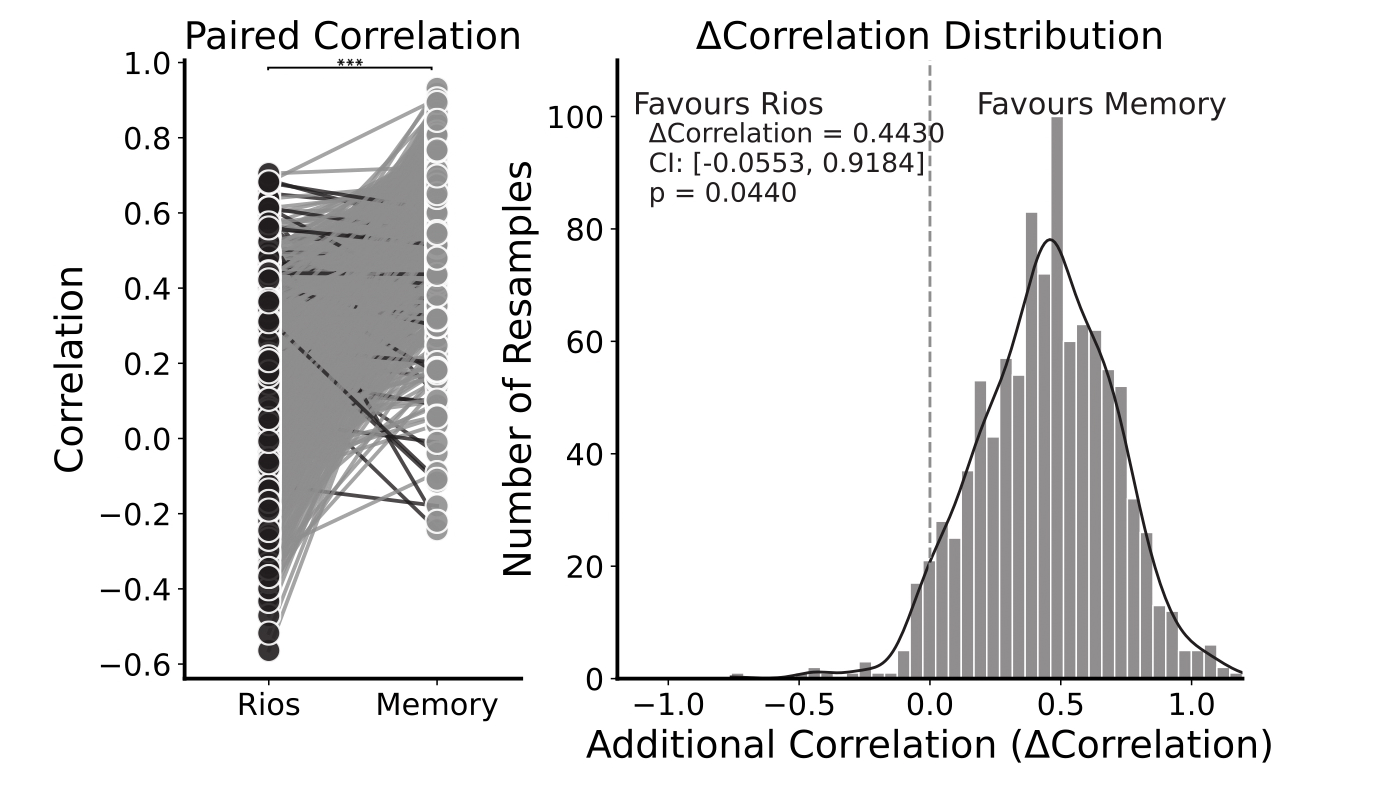
 **Supplementary Figure 9. Spatial Correlation with Convergent Memory Network Correlates More Strongly to Prior Alzheimer Brain Stimulation Trial Outcomes Than Prior Alzheimer Response.** Each study’s stimulation site was seeded to develop a whole-brain connectivity profile and correlated to both the memory target network and a prior network derived by Rios and colleagues meant to predict Alzheimer-specific outcomes after DBS. The spatial correlation of these studies was then correlated with their effect sizes, and the difference was recorded. This process was repeated 1000 times using bootstrapping to estimate the distribution of the true difference. The value of each correlation is plotted on the left, with each bootstrap indicated by a line. The memory target network was found to have significantly higher correlation values than the Rios map (t = 51, p < 0.0001). The distribution of the difference in correlations is shown on the right, demonstrating the memory target network added, on average, 44% additional correlation (p = 0.044).

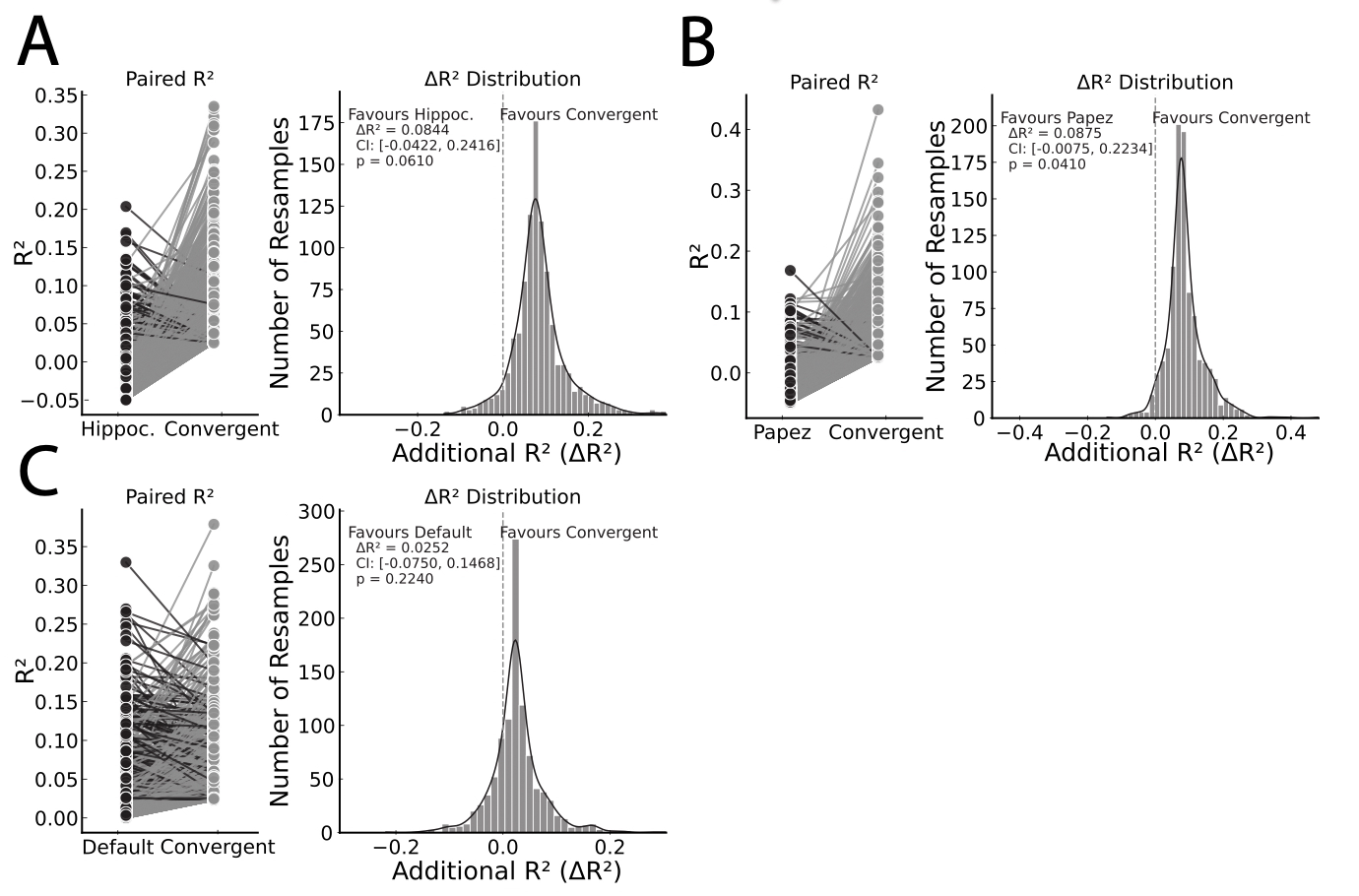
**Supplementary Figure 10. Spatial Correlation with Convergent Memory Network Correlates More Strongly to Prior Alzheimer Brain Stimulation Trial Outcomes Than Other Memory-Related Neuroanatomy.** A) 1,000 bootstrap resamples comparing correlations between outcomes and stimulation-site overlap with the hippocampus versus the convergent memory network demonstrate connectivity to the convergent memory network is more strongly correlated with outcomes (t = 10.7, p <0.0001), explaining on average 8.4% additional variance (p = 0.061). B) 1,000 bootstrap resamples comparing correlations between outcomes and stimulation-site overlap with the Circuit of Papez versus the convergent memory network demonstrate connectivity to the convergent memory network is more strongly correlated with outcomes (t = 17.2, p <0.0001), explaining on average 4.1% additional variance (p = 0.041). C) 1,000 bootstrap resamples comparing correlations between outcomes and stimulation-site overlap with the default mode network versus the convergent memory network demonstrate connectivity to the convergent memory network is more strongly correlated with outcomes (t = 2.4, p = 0.012), explaining on average 2.5% additional variance (p = 0.22).

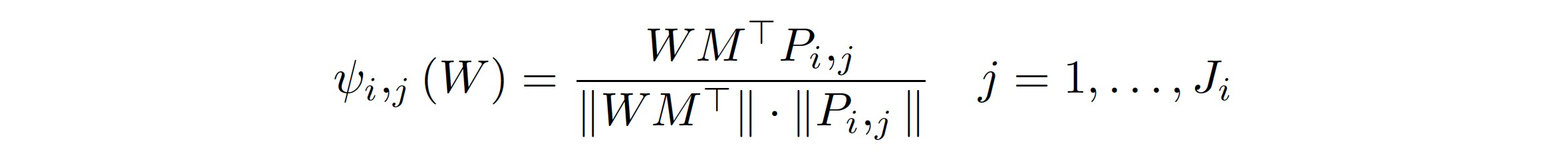
 **Supplementary Equation 1. Definition of similarity between patient connectivity profile and the convergent memory network.** $WM^{\top}$: convergent memory network; $W$*:* weight matrix; $M$*:* fixed group-level memory maps; $P_{i,j}$*:* patient connectivity profile of patient $j$ in dataset $i$; $i$*:* dataset index; $j$: patient index within dataset *i.*

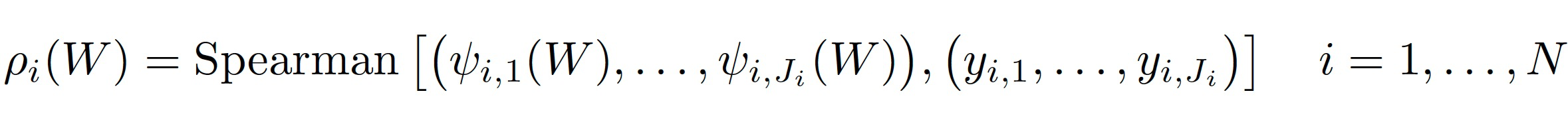
**Supplementary Equation 2. Spearman correlation of similarity scores and outcomes within each dataset.** $\rho_{i}$: dataset-level correlation; $\psi_{i,j}$: similarity (Supplementary Eq. 1) for patient$j$ in dataset $i$; $W$*:* weight matrix; $y_{i,j}$: outcome of patient $j$ in dataset $i$; $i$: dataset index; $j$: patient index within dataset $i$.

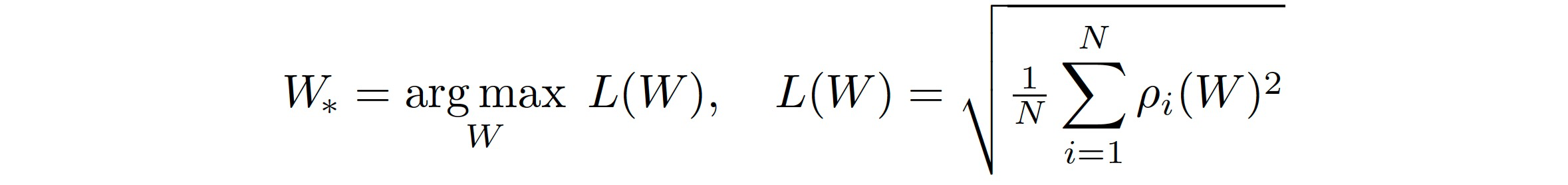
**Supplementary Equation 3. Optimization objective to determine the weight matrix** $\boldsymbol{W}_{\boldsymbol{*}}$**, maximizing the root mean squared spearman rho across datasets.** $W_{*}$: optimized weight matrix; $L(W)$: Root Mean Squared Spearman Rho correlation objective; $\rho_{i}$: dataset-level spearman correlation (Supplementary Eq. 2); $i$: dataset index.

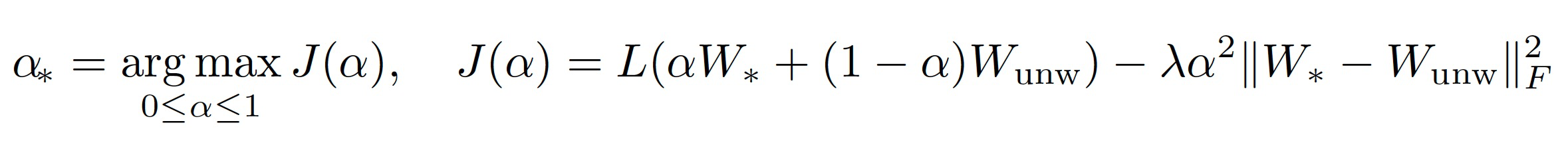
**Supplementary Equation 4. Optimization objective to determine blending factor** $\boldsymbol{\alpha}_{\boldsymbol{*}}$**, balancing optimized weighted matrix and unweighted baseline.** $\alpha_{*}$: optimized blending factor; $W_{*}$: optimized weight matrix (Supplementary Eq. 3); $W$*:* weight matrix; $W_{unw}$*:* unweighted weight matrix; $\lambda$: regularization factor controlling deviation from baseline unweighted matrix; $\left\| \left. \cdot\right\|_{F} \right.$: Frobenius norm.
